# Lesion-guided stereotactic radiofrequency thermocoagulation for drug-resistant focal epilepsy: preliminary multi-center report from Japan

**DOI:** 10.1101/2025.05.20.25327981

**Authors:** Masaki Iwasaki, Takahiro Hayashi, Keiya Iijima, Yuiko Kimura, Naoki Ikegaya, Yutaro Takayama, Masaki Sonoda, Takashi Morishita, Koichi Hagiwara, Masafumi Fukuda, Tomotaka Ishizaki, Satoshi Maesawa

**Author notes:** **Corresponding Author:** Masaki Iwasaki, MD, PhD, Department of Neurosurgery, National Center Hospital, National Center of Neurology and Psychiatry, 4-1-1 Ogawahigashicho, Kodaira, Tokyo 187-8551, Japan.

## Abstract

**Introduction:** Although lesion-guided stereotactic radiofrequency thermocoagulation (RFTC) is being increasingly employed, data regarding its clinical outcomes and patient selection criteria remain limited. This study aims to elucidate the current status of RFTC for epilepsy in a multi-center Japanese cohort.

**Methods:** This retrospective study included 23 patients who underwent lesion-guided RFTC for drug-resistant focal epilepsy between January 2021 and April 2024. Pre- and postoperative clinical data were collected and analyzed in relation to postoperative seizure outcomes.

**Results:** The median age at surgery was 16 years, with a median follow-up of 27 months. The most frequent etiology was focal cortical dysplasia (60.9%). Surgical planning was primarily based on MRI and FDG-PET findings, supplemented by stereo-electroencephalography (SEEG) in most cases. The median number of ablations per patient was 23, ranging from 5 to 51. The treatment area included the insulo-opercular cortices in 11 patients and the medial temporal lobe in 5 patients. No surgical complications occurred, although transient and permanent neurological deficits were observed in 34.8% and 13.0% of patients, respectively. Seizure freedom was achieved in 59.1% of patients at 1 year and 34.8% at the last follow-up. Prior epilepsy surgery was significantly associated with poorer seizure outcomes (p = 0.02). No other preoperative factors demonstrated a significant association with seizure freedom.

**Conclusion:** Lesion-guided RFTC appears to be a safe and effective, less invasive surgical option for selected patients with drug-resistant focal epilepsy, particularly those with deep-seated lesions or those involving eloquent cortex. While short-term seizure control is encouraging, long-term outcomes remain suboptimal, underscoring the need for improved patient selection and standardized treatment protocols.

## Introduction

Epilepsy surgery has been accepted as a treatment option for individuals with drug-resistant focal epilepsy.^1, 2)^ Conventional epilepsy surgeries, such as resective procedures, have demonstrated substantial efficacy in achieving seizure control and enhancing patients’ quality of life.^3)^ However, these surgeries are often invasive and carry significant risks, particularly when lesions are located in or near eloquent cortical regions. When lesions are located in deep areas, such as the hippocampus and insula, surgical access may cause collateral damage to the overlying cortices.^4, 5)^ This can lead to neurological deficits and reduced functional outcomes. Thus, there is continuous motivation to develop safer and less invasive surgical alternatives.

In recent years, there has been a growing interest in minimally invasive surgical approaches, particularly stereotactic ablation techniques.^6-8)^ These methods, including stereotactic radiofrequency thermocoagulation (RFTC) and laser interstitial thermal therapy (LITT), allow precise destruction of epileptogenic tissue while preserving adjacent brain structures. Stereotactic ablations may offer advantages such as reduced morbidity, shorter hospitalization, and faster recovery times, which have increased their attractiveness as initial surgical options. Stereotactic ablation techniques have shown comparable outcomes to conventional surgery in selected patient groups, further justifying their growing clinical adoption.^9)^

The use of medical devices is often limited by approval status and cost. In Japan, LITT is currently unavailable, and therefore, interest in RFTC as an alternative less invasive option is increasing.^10)^ RFTC has largely been established as a treatment option for hypothalamic hamartoma,^11)^ and its application to other types of epileptogenic lesions has only recently begun. Although RFTC is being increasingly employed, data regarding its clinical outcomes and patient selection criteria remain limited. This study aims to elucidate the current status of RFTC in Japan by examining its clinical application, evaluating postoperative seizure outcomes, identifying potential prognostic factors, and highlighting the challenges to this treatment approach.

## Materials and Methods

This was a multi-center, retrospective cohort study approved by the Ethics Committee of the National Center of Neurology and Psychiatry (Number B2024-116) in accordance with the ethical standards of the 1964 Declaration of Helsinki and its later amendments or comparable ethical standards. The requirement for written informed consent was waived because of the retrospective study design.

### Study population

This study included patients with focal epilepsy who underwent stereotactic radiofrequency thermocoagulation (RFTC) aimed at controlling drug-resistant seizures and had a minimum of 1 year of follow-up. Patients with hypothalamic hamartoma or cerebellar hamartoma were excluded. Note that the RFTC here was not SEEG-guided but was performed as a separate procedure from SEEG. SEEG-guided RFTC was not technically available for clinical purposes at the time of this study in Japan because depth electrodes were not approved for delivering radiofrequency thermocoagulation.

### Data collection

The following data were retrospectively collected from medical records: basic patient demographics including birthdate, sex, past medical history, and family history of epilepsy or seizures, age at onset of epilepsy, seizure frequency at the time of surgery, anti-seizure medications prescribed at the time of surgery and in the past, International League Against Epilepsy (ILAE) classification of epilepsy,^12)^ seizures,^13)^ and epilepsy syndrome,^14)^ etiology of epilepsy, presence of intellectual and/or developmental impairments, pre-operative findings of long-term video-electroencephalogram monitoring, brain magnetic resonance imaging (MRI), fluoro-deoxy-glucose positron emission tomography (FDG-PET), ictal single photon emission computed tomography (SPECT), magnetoencephalography (MEG), and intracranial EEG evaluation, methods of RFTC surgery including the stereotactic devices and planning software used, targeted temperature and duration of ablation, number of insertions and ablations, operation duration, counter-measures for post-operative thermal edema, postoperative surgical and neurological complications, date of seizure recurrence, and post-operative seizure outcomes at 1 year and at the last follow-up.

Postoperative seizure outcome was assessed using the ILAE classification.^15)^ For patients who underwent repeat epilepsy surgery, the seizure outcome was assessed at the time of repeat surgery. Information was obtained on which pre-operative findings were used for planning ablations. The indicators of ablation planning were divided into seizure onset zone (SOZ) in the intracranial EEG study, MRI lesion, FDG-PET hypometabolism, ictal SPECT, MEG spike mapping, semiological features, and seizure induction test with electrical stimulation.

### Statistical analysis

The patient demographics were characterized using descriptive statistics. Data were presented with medians with ranges, proportions, and frequencies. Univariate comparisons were performed to analyze the relationship between preoperative factors and seizure freedom at one year. Fisher’s exact test was used for categorical variables, and Student’s t-test was used for continuous variables. The Kaplan–Meier survival curve was calculated for time to seizure recurrence after surgery. R version 4.1.0 and above (The R Foundation for Statistical Computing) was used for statistical analysis. Statistical significance was accepted at p < 0.05.

## Results

### Patient clinical characteristics

A total of 23 patients, including 15 males, were enrolled in this study. RFTC was performed during the period between January 2021 and April 2024. Table 1 lists the preoperative clinical characteristics of patients. Median ages at epilepsy onset and surgery were 5 years (0–36) and 16 years (2–56), respectively. Six patients had a previous history of epilepsy surgery and underwent RFTC as repeat epilepsy surgery. All patients had drug-resistant seizures. Twelve patients had daily seizures, and the most common seizure type was focal impaired awareness seizures (n=15). The median number of previously failed anti-seizure medications was 5 (2–11). Intellectual impairment or developmental delay was noted in 9 patients. The most frequent etiology was focal cortical dysplasia (FCD), seen in 14 patients (60.9%). Among them, 4 patients had bottom-of-sulcus (BOS) type dysplasia. Two patients suffered from post-encephalitis epilepsy, and 3 patients had unknown causes of epilepsy. These 5 patients showed no causative structural abnormalities in MRI, although 3 patients had scarring lesions associated with previous epilepsy surgery. The other 18 patients were positive for epileptogenic MRI lesions.

**Table 1.** Preoperative characteristics.

| Case | Sex | Onset of epilepsy* [Y] | Age at surgery* [Y] | Previous history of epilepsy surgery | N of previously failed anti-seizure medications | Sz freq | Seizure types | Etiology of epilepsy | Ictal EEG | Interictal EEG | MRI lesion location | Additional presurgical evaluations |  |  |  |
| --- | --- | --- | --- | --- | --- | --- | --- | --- | --- | --- | --- | --- | --- | --- | --- |
|  |  |  |  |  |  |  |  |  |  |  |  | PET | Ictal SPECT | MEG | Intracranial EEG |
| 1 | F | 0-5 | 0-5 |  | 5 | daily | FIAS | FCD | L T | none | Lt temporo-parietal operculum | y | y | y | SEEG |
| 2 | F | 0-5 | 6-10 |  | 5 | daily | FAS | FCD | L P | L P | Lt parietal operculum | y |  | y | SEEG |
| 3 | M | 6-10 | 21-25 |  | 6 | weekly | FAS | FCD | R F | R F | Rt bottom of the precentral sulcus | y |  | y | not done |
| 4 | F | 0-5 | 6-10 |  | 5 | daily | FBTCS | FCD | L C | L C | Lt frontal operculum | y |  | y | SEEG |
| 5 | F | 6-10 | 31-35 |  | 11 | daily | FAS | FCD | Vertex | Vertex | Rt bottom of the superior frontal sulcus | y |  | y | not done |
| 6 | M | 16-20 | 16-20 |  | 2 | weekly | FIAS | FCD | R T | R aT | Rt lingual gyrus | y |  |  | SEEG |
| 7 | F | 11-15 | 16-20 |  | 3 | daily | FBTCS | FCD | L F | none | Lt superior frontal gyrus | y |  |  | SEEG |
| 8 | M | 0-5 | 11-15 |  | 4 | weekly | FAS | FCD | R C | R C | Rt bottom of the central sulcus | y | y | y | SEEG |
| 9 | F | 6-10 | 11-15 |  | 5 | monthly | FAS | FCD | L T | L T | Lt supra-marginal gyrus | y |  | y | SEEG |
| 10 | M | 0-5 | 6-10 | y | 3 | daily | FAS | FCD | L hemispheric | L C | Lt insula to frontal operculum | y | y | y | SEEG |
| 11 | M | 0-5 | 0-5 |  | 10 | daily | FIAS | FCD | L aT | L T, R F-C-P | Lt insula | y | y | y | SEEG |
| 12 | M | 0-5 | 11-15 |  | 8 | daily | FIAS | FCD | L hemispheric | L T | Lt insula - temporo-parietal operculum | y | y | y | SEEG |
| 13 | M | 0-5 | 11-15 | y | 8 | daily | FIAS | FCD | R F-C-T | none | Rt insula | y |  | y | SEEG |
| 14 | M | 0-5 | 0-5 |  | 7 | daily | FIAS | FCD | R hemispheric | R aT | Rt anterior temporal, amygdala, and insula | y |  | y | SEEG |
| 15 | F | 16-20 | 51-55 | y | 9 | weekly | FIAS | Periventricular nodular heterotopia | Blt T | Blt T | Blt periventricle | y |  | y | Subdural electrodes |
| 16 | M | 11-15 | 16-20 |  | 8 | monthly | FIAS | Polymicrogyria | R T | R T | Rt temporo-parietal | y |  | y | SEEG |
| 17 | M | 26-30 | 46-50 |  | 3 | monthly | FIAS | HS | R T | R aT | Rt hippocampus | y |  | y | not done |
| 18 | M | 11-15 | 36-40 |  | 3 | daily | FIAS | HS | L T | none | Lt medial temporal | y |  |  | SEEG |
| 19 | M | 36-40 | 56-60 | y | 5 | weekly | FIAS | Infectious | L aT | L aT | no lesion |  | y | y | SEEG |
| 20 | M | 0-5 | 16-20 | y | 7 | weekly | FIAS | Infectious | R F | R C | no lesion | y |  |  | SEEG |
| 21 | M | 0-5 | 6-10 |  | 9 | daily | FIAS | unknown | L hemispheric | Generalized | no lesion | y | y | y | SEEG |
| 22 | M | 26-30 | 41-45 |  | 3 | monthly | FIAS | unknown | R T | R aT | no lesion |  |  | y | not done |
| 23 | F | 0-5 | 31-35 | y | 3 | monthly | FIAS | unknown | L aT | L aT | no lesion |  |  | y | SEEG |

\* described by age range
F = female; M = male; FIAS = focal impaired awareness seizure; FAS = focal aware seizure; FBTCS = focal to bilateral tonic-clonic seizure; FCD = focal cortical dysplasia; HS = hippocampal sclerosis; T = temporal; P = parietal; F = frontal; C = central; aT = anterior temporal; SEEG = stereo-electroencephalography

FDG-PET, ictal SPECT, and MEG spike mapping were performed in 20, 7, and 19 patients, respectively. Epilepsy-related local metabolic changes were found in all patients who underwent FDG-PET. Ictal hyperperfusion was observed in 6 of the 7 patients (85.7%) who underwent ictal SPECT. Magnetic source imaging was available in 13 cases (68.4%), while epileptiform discharges were not detected in the other 6 cases in MEG. Invasive evaluation with intracranial electrode implantation was performed in 19 patients (82.6%). All except one case was evaluated with stereo-electroencephalography (SEEG). At least one epileptic seizure was recorded in all cases.

### Surgical planning and procedures

Surgical planning and RFTC procedures are summarized in Table 2. Ablation was primarily planned based on MRI/PET lesions and the SOZ identified through intracranial EEG. A representative case is shown in Figure 1. SOZ was utilized in the planning for 18 out of 19 patients (94.7%). MRI findings were utilized in 20 patients (87.0%). PET abnormalities were used in 85.0% (n=17) of patients when available. Notably, ablation was planned solely on MRI/PET findings without intracranial EEG in 4 patients, including 3 individuals with BOS-FCD. During planning, ictal SPECT and MEG spike maps were used in 57% (4 of 7 patients) and 36.8% (7 of 19 patients) of cases, respectively. Semiological features and seizure induction testing with electrical stimulation were also utilized in 19 and 14 patients, respectively.

**Table 2.** Surgical planning and procedures.

| Case | Indicators of treatment planning |  |  |  |  |  |  | Side of surgery | Cranial opening methods | N of trajectories | N of ablations | Lesioning area | Operation time [min] |
| --- | --- | --- | --- | --- | --- | --- | --- | --- | --- | --- | --- | --- | --- |
|  | Seizure onset zone | MRI lesion | PET lesion | Ictal SPECT | MEG | Semiology | Sz induction with electrical stimulation |  |  |  |  |  |  |
| 1 | y | y | y |  |  | y | y | Lt | Craniotomy | 13 | 48 | Temporo-parietal operculum | 393 |
| 2 | y | y | y |  |  | y | y | Lt | Craniotomy | 14 | 33 | Parietal operculum | 358 |
| 3 |  | y | y |  |  | y |  | Rt | Craniotomy | 12 | 31 | BOS lesion (precentral sulcus) | 247 |
| 4 |  | y |  |  |  |  |  | Lt | Craniotomy | 13 | 20 | BOS lesion (frontal operculum) | 452 |
| 5 |  | y | y |  |  | y |  | Rt | Craniotomy | 9 | 15 | BOS lesion (superior frontal sulcus) | 158 |
| 6 | y | y | y |  |  | y | y | Rt | Burr holes | 9 | 33 | Lingual gyrus | 307 |
| 7 | y | y | y |  |  | y | y | Lt | Burr holes | 6 | 13 | Cingulate gyrus | 215 |
| 8 | y | y | y |  | y | y | y | Lt | Craniotomy | 9 | 22 | BOS lesion (central sulcus) | 218 |
| 9 | y | y | y |  |  | y | y | Lt | Burr holes | 18 | 51 | Supra-marginal gyrus | 537 |
| 10 | y | y | y | y | y | y | y | Lt | Craniotomy | 9 | 23 | Insula | 322 |
| 11 | y | y | y | y |  | y | y | Lt | Craniotomy | 11 | 31 | Insula | 403 |
| 12 | y | y | y | y | y | y | y | Lt | Craniotomy | 14 | 39 | Insula, Temporo-parietal operculum | 455 |
| 13 | y | y | y |  |  | y | y | Lt | Craniotomy | 8 | 29 | Insula | 264 |
| 14 | y | y | y |  |  | y | y | Rt | Craniotomy | 6 | 21 | Insula | 254 |
| 15 | y | y |  |  |  |  |  | Rt | Craniotomy | 7 | 20 | Periventricular lesion | 164 |
| 16 | y |  |  |  |  |  | y | Rt | Craniotomy | 9 | 22 | Temporo-perietal lesion | 256 |
| 17 |  | y | y |  | y | y |  | Rt | Burr holes | 4 | 20 | Medial temporal | 163 |
| 18 | y | y | y | y |  |  |  | Lt | Twist holes | 5 | 8 | Hippocampus | 198 |
| 19 | y | y |  | y | y | y |  | Lt | Burr holes | 10 | 35 | Insula, Temporal lobe | 285 |
| 20 | y | y | y |  |  | y | y | Rt | Craniotomy | 8 | 24 | Insula | 199 |
| 21 | y | y | y |  |  | y | y | Lt | Craniotomy | 7 | 21 | Insula | 291 |
| 22 |  |  |  |  | y | y |  | Rt | Burr holes | 4 | 24 | Medial temporal | 182 |
| 23 | y |  |  |  | y | y |  | Lt | Burr holes | 6 | 21 | Medial temporal | 162 |

BOS: bottom-of -sulcus

**Figure 1.**
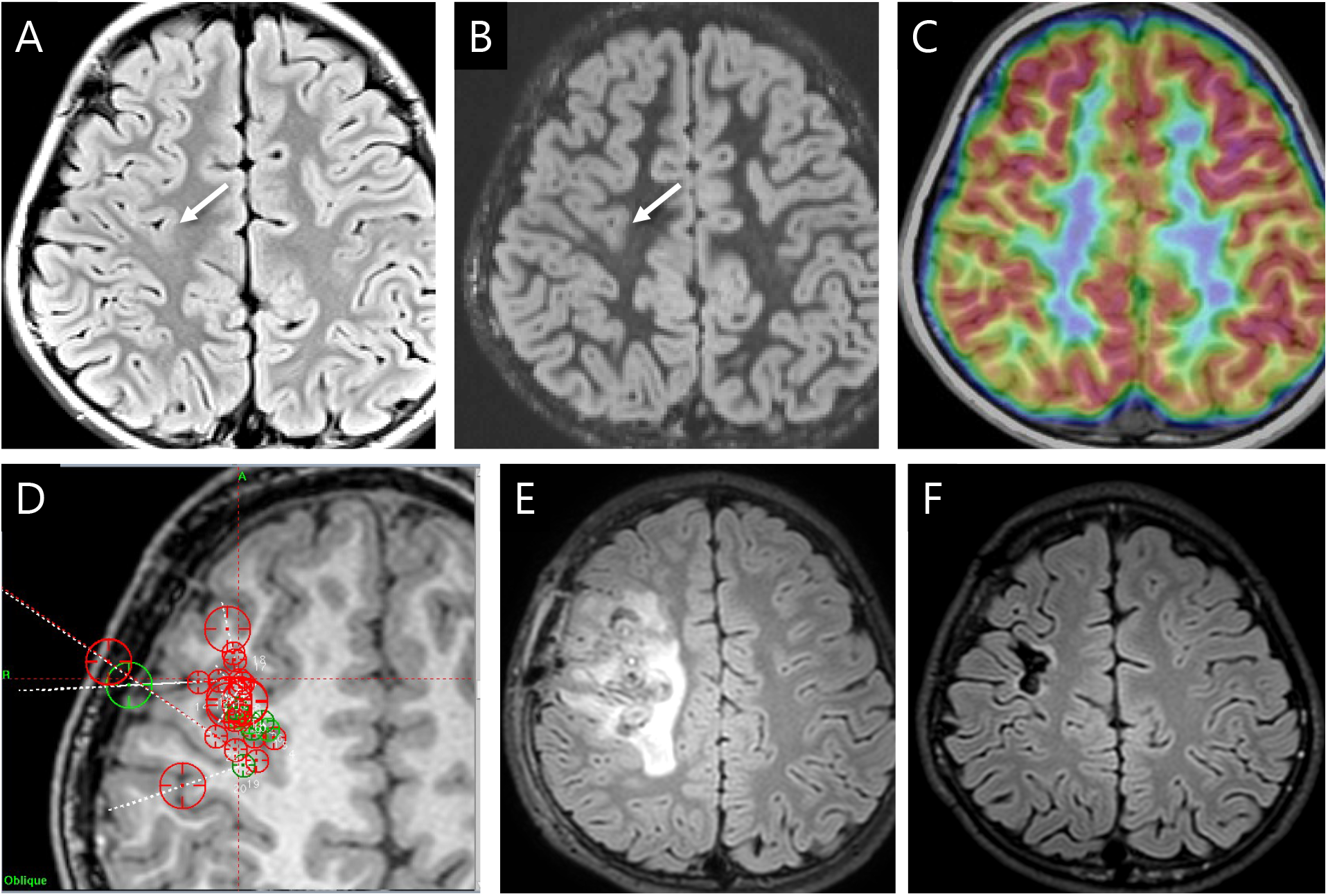
A case of lesion-guided stereotactic radiofrequency thermocoagulation for bottom-of-sulcus type dysplasia in the right central region. A teenage boy suffered epileptic seizures for 6 years. His seizures were characterized by abnormal sensation in the left face and upper limb, followed by eye deviation to the left and tonic-clonic activity in the left upper limb. Despite multiple anti-seizure medications including carbamazepine, levetiracetam, lamotrigine, and perampanel, he continued to have weekly seizures. Fluid-attenuated inversion recovery (FLAIR) image (A) and double inversion recovery image (B) showed a blurred boundary between cortex and white matter at the bottom of the abnormal sulcus, extending to the central sulcus. Fluoro-deoxy-glucose positron emission tomography (C) showed mild glucose hypometabolism in the right pre- and post-central gyri. Focal cortical dysplasia was suspected after comprehensive presurgical evaluations including long-term video-electroencephalography monitoring, and subsequent stereo-electroencephalography identified seizure onset from the suspected lesion. Electrical stimulation of the lesion successfully reproduced his habitual seizures, further supporting localization. Stereotactic radiofrequency thermocoagulation was planned to target the sulcus bottom with multiple trajectories (D). A total of 22 ablations were performed using 9 trajectories. Acute postoperative FLAIR image (E) showed marked thermal edema around the ablation site, which resolved completely 2 years after surgery (F). The patient had rare focal motor seizures, managed with medication adjustments. He was left with moderate motor weakness and mild sensory disturbance confined to the left-hand digits.

Leksell Surgiplan® (Elekta AB, Stockholm) was used to plan ablations in 20 cases. Brainlab iPlan or Elements® (Brainlab AG, Munich) was used in the remaining 3 cases. All frame-based stereotactic procedures were performed using the Leksell Stereotactic System® (Elekta, Stockholm). Two procedures were guided by a stereotactic robotic system (Neuromate®, Renishaw plc., Gloucestershire). Surgery was performed under craniotomy in 15 patients (65.2%). The targeted temperature and duration of thermocoagulation were set at 74 °C for 60 seconds in 20 patients, which is known to produce a 5 mm diameter thermal lesion. Lower settings (70 °C for 60 seconds, 64 °C for 44 seconds) were used in 3 patients. Anti-inflammatory drugs, mainly dexamethasone, were used perioperatively in all but one patient to reduce thermal edema.

The median numbers of trajectories and ablations were 9 (4-18) and 23 (5-51), respectively, indicating that an average of 2.86 lesions were created per insertion. Eleven patients underwent ablation of the insulo-opercular cortices, and 5 underwent ablation of the medial temporal region. The median operation duration was 256 minutes (158-537).

### Postoperative outcomes

Postoperative outcomes are summarized in Table 3. No surgical complications occurred. Transient and permanent neurological sequelae were observed in 8 and 3 patients, respectively. All were expected from the treatment site. Patient 8 was treated for FCD located at the bottom of the central sulcus at the level of the upper extremity, which was anticipated to result in left upper limb motor weakness; 2 years later, the patient was left with moderate paresis and mild sensory disturbance confined to the hand digits (Figure 1).

**Table 3.** Postoperative outcomes.

| Case | Transient neurological deficits | Permanent neurological deficits | Seizure outcome at 1 year* | Seizure outcome at last follow-up* | Follow-up period [M] | Repeat epilepsy surgery |
| --- | --- | --- | --- | --- | --- | --- |
| 1 |  |  | 1 | 1 | 39 |  |
| 2 |  |  | 1 | 1 | 23 |  |
| 3 | Lt face and hand weakness |  | 1 | 1 | 13 |  |
| 4 | Rt oro-facial motor weakness |  | 1 | 1 | 50 |  |
| 5 | Lt hemiparesis |  | 1 | 1 | 12 |  |
| 6 |  | Lt upper quadrantanopsia | 1 | 1 | 13 |  |
| 7 | Rt hemiparesis |  | 1 | 2 | 30 |  |
| 8 | Lt face and upper limb weakness | Lt upper limb sensori-motor disturbance | 1 | 3 | 25 |  |
| 9 | Aphasia |  | 3 | 3 | 14 |  |
| 10 |  |  | 4 | 4 | 32 |  |
| 11 |  |  | 1 | 5 | 47 |  |
| 12 |  |  | 5 | 5 | 37 |  |
| 13 |  |  | 5 | 5 | 12 |  |
| 14 | Lt hemiparesis |  |  | 5 | 37 | Temporal lobectomy |
| 15 |  | Alexia and agraphia for Kanji characters | 4 | 4 | 21 |  |
| 16 |  |  | 5 | 5 | 41 | Corticectomy |
| 17 |  |  | 1 | 1 | 27 |  |
| 18 |  |  | 5 | 5 | 40 | Temporal lobectomy |
| 19 |  |  | 2 | 2 | 49 |  |
| 20 |  |  | 4 | 4 | 26 |  |
| 21 |  |  | 1 | 1 | 27 |  |
| 22 |  |  | 3 | 3 | 31 |  |
| 23 | Rt visual field deficit |  | 5 | 5 | 23 |  |

\* International League Against Epilepsy classification (ref. 15)

Seizure freedom was achieved in 13 patients at one year after surgery (59.1%) and in 8 patients at the last follow-up (34.8%). Significant seizure reduction (class 4 or better) was observed in 17 patients at 1 year (77.3%) and in 16 patients at the last follow-up (69.6%). The median follow-up period was 27 months (12-50). Three patients subsequently underwent repeat epilepsy surgery for residual seizures. One patient underwent temporal lobectomy 7 months postoperatively, another at 30 months, and a third individual underwent cortical resection 23 months postoperatively. The final seizure outcome was judged as class 5 at the time of reoperation in those patients. The median follow-up to the final seizure outcome evaluation was 26 months (7-50).

Previous history of epilepsy surgery was associated with a lower likelihood of seizure freedom (p=0.02). No significant associations were found for other preoperative factors, including sex (p=0.38), side of surgery (p=0.66), etiology of FCD (p=0.38), insulo-opercular lesioning (p=0.67), daily seizures (p=0.39), and intellectual or developmental impairment (p=0.66). There were no differences in age at surgery (p=0.30), duration of epilepsy (p=0.49), and number of ablations (p=0.25) between patients who achieved seizure freedom and those who did not.

The Kaplan–Meier survival curve for postoperative time to seizure recurrence is shown in Figure 2. The probability of being seizure-free was 60.9% at 1 year after surgery in the total cohort (95% CI: 38.3–77.4%). These probabilities were 70.6% in patients with a prior history of epilepsy surgery (n=17, 95% CI: 43.2–86.6%) and 33.3% in those without (n=6, 95% CI: 4.61–67.6%), with no statistical difference (p=0.141, log-rank test).

**Figure 2.**
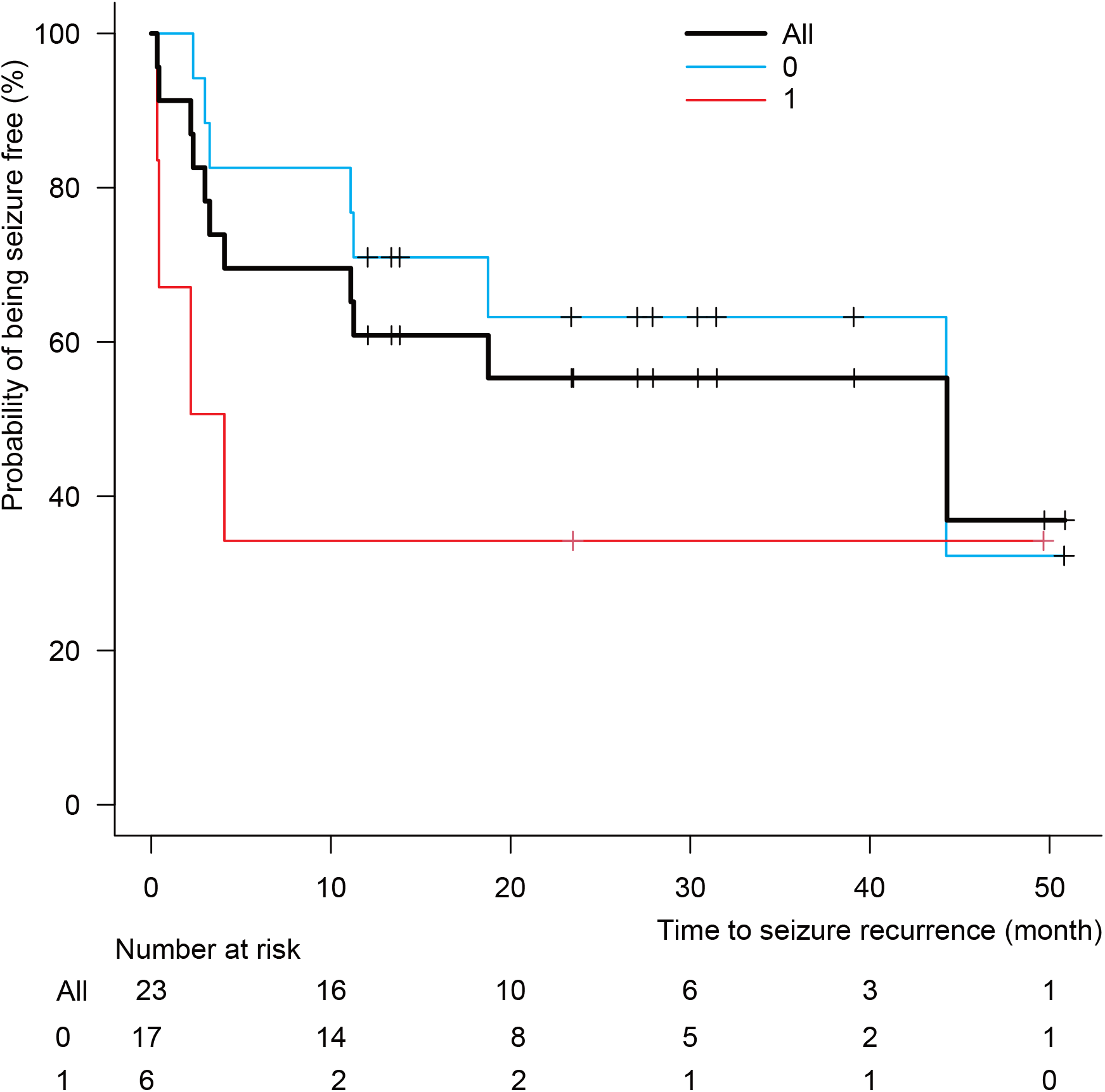
Kaplan–Meier curve of the time to seizure recurrence. The probability of seizure freedom was 60.9% at 12 months after surgery (black). Time to recurrence tended to be shorter in individuals with a previous history of epilepsy surgery (red) compared with those without (blue), but the difference was not statistically significant.

## Discussion

This study provides preliminary support for the utility of RFTC in treating drug-resistant focal epilepsy, with a 60.9% probability of remaining seizure-free at one year. The result aligns with previous reports and underscores RFTC as a promising option. However, further studies with larger cohorts are necessary to elucidate long-term outcomes. In this study, most treatments were planned based on MRI and FDG-PET lesions, often supported by SEEG findings, which should be referred to as “lesion-guided” RFTC. RFTC was selectively performed for treating insulo-opercular lesions, medial temporal lesions, and other eloquent area lesions. Our lesion-guided RFTC was performed as a separate procedure from SEEG, allowing for more adequate coverage for coagulation. Lesion-guided RFTC appears superior to SEEG-guided RFTC in terms of seizure control,^16)^ but slightly inferior to conventional resective epilepsy surgery. Its low invasiveness allows for repeated procedures in cases of seizure recurrence, which may improve seizure outcomes. We believe lesion-guided RFTC can be effectively used for curative treatment of epilepsy in carefully selected patients.

Our seizure outcomes are comparable to previous studies evaluating RFTC and similar minimally invasive treatments such as LITT. Seizure-free outcomes were reported in 53% of patients in a report on RFTC for insular epilepsy^17)^ and in 5 of 7 patients (71.4%) in a report on BOS-FCD.^18)^ A recent systematic review revealed that “monopolar” RFTC was associated with a 50.0% rate of Engel I/II outcomes (n = 166);^19)^ the monopolar RFTC here was delivered with a coagulating probe in a staged fashion, similar to our cases. A recent meta-analysis of LITT studies showed a 57.8% seizure-free rate, with mesial temporal sclerosis and lesional MRI being positive predictors.^6)^ At the last follow-up, fewer than 40% of patients had maintained seizure freedom in our study, suggesting that the long-term outcome remains suboptimal. RFTC remains an alternative to open surgery, but better patient selection and treatment planning should be investigated.

RFTC is appropriate for more selective treatment of deep-seated lesions. The insula, medial temporal lobe, periventricular heterotopia, and hypothalamic hamartoma have previously been reported as representative treatment sites for stereotactic ablations.^6, 10, 11, 17, 19, 20)^ RFTC may be the first option for these regions because of the technical difficulties and risks involved in surgical approaches. BOS dysplasia is another suitable indication for RFTC. A recent study showed that postoperative seizure-free rates were comparable between LITT and open surgery, but the removal volume was smaller with LITT, indicating that stereotactic ablation provides good efficacy with less invasiveness. Considering that BOS-FCD often occurs around the central sulcus,^21, 22)^ RFTC is likely to be applied in many situations as a treatment associated with fewer complications.

Patient selection is important for good outcomes. Previous epilepsy surgery had a negative impact on postoperative seizure freedom in this study. This finding highlights the challenges in reoperative epilepsy surgery, possibly reflecting more complex epilepsy networks in individuals who continue to have seizures after surgical interventions. Treatment volume, expressed as the number of ablations, had no apparent association with seizure outcomes in this study. It is a rule of thumb in epilepsy surgery that the larger the treatment volume, the better the seizure outcome. This holds true in general, including in the collected data of RFTC.^19)^ However, Mullatti et al. reported that a better outcome was significantly associated with a low volume of RFTC in their cases with insular epilepsy.^17)^ This indicates that accurate localization of a small, circumscribed epileptogenic lesion is critical in RFTC.

Stereotactic RFTC can be performed relatively safely. No significant surgical complications were noted in this small study. The relatively high incidence of transient neurological deficits (34.8%) was anticipated and related directly to the anatomical targets chosen for ablation. Postoperative thermal edema may have exacerbated transient neurological symptoms. Measures to reduce postoperative edema are important. Permanent deficits, while less frequent, underline the importance of careful patient selection and risk-benefit evaluation, particularly for lesions near the eloquent cortex. Our safety results are comparable to previous reports.^19)^

Limitations of this study include its retrospective design, small patient sample, and variable follow-up durations. Prospective studies with larger cohorts and standardized protocols are warranted to confirm these preliminary findings and refine patient selection criteria. The lack of significant associations with many preoperative factors is likely due to small sample sizes. Further research with larger cohorts is necessary to better define the predictors of successful outcomes. Another limitation is the lack of standardization in surgical planning methods and coagulation settings. The variability in these procedures may introduce bias and affect the consistency of the results. Future studies should aim to implement standardized protocols to minimize these discrepancies and improve the reliability of the findings.

In conclusion, our preliminary results support RFTC as an effective surgical option for selected individuals with drug-resistant focal epilepsy. Identification of clear preoperative predictors for long-term seizure freedom remains crucial and necessitates further investigations.

## Acknowledgment

This study was supported in part by Grants-in-Aid for Scientific Research (KAKENHI, grant number JP22K09296) from the Japan Society for the Promotion of Science (JSPS), by the Japan Agency for Medical Research and Development (AMED) under grant numbers JP24wm0625407, JP24wm0425095 and JP24ek0109764, and by the Intramural Research Grant (5-4: Research on sustainable and advanced epilepsy care and database with telemedicine) for Neurological and Psychiatric Disorders of the National Center of Neurology and Psychiatry.

## Conflicts of Interest Disclosure

All authors have no conflict of interest and have registered online self-reported COI disclosure statement forms through the Japan Neurosurgical Society members’ website. We would like to thank Editage (www.editage.jp) for English language editing.

## Data Availability Statement

All data produced in the present study are available upon reasonable request to the authors

## Notes

### Competing Interest Statement

The authors have declared no competing interest.

### Author Declarations

Ethics Committee of the National Center of Neurology and Psychiatry gave ethical approval of this work.

